# *ASXL1-*Mutant Clonal Hematopoiesis is Associated with Calcific Aortic Valve Disease and Promotes Valvular Calcification *In Vitro*

**DOI:** 10.64898/2026.09.23.26363798

**Authors:** Aeron M. Small, Liying Xue, Shinsuke Itoh, Cesar De Jeronimo Diaz, Yuto Nakamura, Niekbachsh Mohammadnia, Huajun Liao, Yota Maekawa, Linke Li, Sasha A. Singh, Taku Kasai, Luisa Weiss, Mesbah Uddin, Art Schuermans, Spencer Flynn, Elby MacKenzie, James C. Engert, George Thanassoulis, Peter Libby, Amil M. Shah, Christie M. Ballantyne, Pradeep Natarajan, Trevor P. Fidler, Elena Aikawa, Michael C. Honigberg

## Abstract

**Background and Aims:** Calcific aortic valve disease (CAVD) is a common valvular heart condition and has no effective medical therapy. Clonal hematopoiesis of indeterminate potential (CHIP), characterized by the acquisition of somatic mutations in hematopoietic stem cells which provides a selection advantage, has been associated with incident aortic stenosis (AS), particularly when driven by *TET2* mutations.

**Methods:** We evaluated the association of CHIP and gene-specific CHIP subtypes with AS in the UK Biobank (UKB) and with aortic valve hemodynamics in the Atherosclerosis Risk in Communities (ARIC) study to prioritize gene driver mutations for mechanistic study. We then generated human *ASXL1*-mutant THP-1 derived macrophage like cells and evaluated the ability of THP-1 conditioned media to promote a calcific response in human valvular interstitial cells (VICs). We additionally performed mass spectrometry proteomics to characterize the *ASXL1-*mutant THP-1 macrophage secretome. Therapeutically relevant inflammatory pathways were inhibited with anakinra (interleukin [IL]-1) or tocilizumab (IL-6).

**Results:** Among 449,109 individuals in the UKB with whole exome sequencing, large CHIP clones and non-*DNMT3A* CHIP subtypes (e.g., *TET2*, *ASXL1*, and *JAK2* CHIP) were independently associated with risk of incident AS. Among 1,963 individuals in ARIC with CHIP sequencing and echocardiographic data, only *ASXL1* CHIP was associated with worse aortic valve hemodynamics. *In vitro, ASXL1*-mutant human THP-1-derived macrophage-like cells had increased AIM2 inflammasome activation. THP-1 media promoted VIC calcification, which was accelerated in *ASXL1* mutant macrophages. Proteomic profiling identified inflammatory proteins linked to valvular calcification. Accelerated VIC calcification was reduced by IL-1 inhibition by anakinra or IL-6 inhibition by tocilizumab.

**Conclusions:** Integrating human cohort data with experimental evidence using *ASXL1*-mutant human macrophage-like cells and human valve cells, these findings identify *ASXL1*-mutant CHIP as a distinct inflammatory subtype associated with valvular calcification and support IL-1 and IL-6 signaling as candidate therapeutic pathways for future investigation in CAVD.

## Introduction

Aortic stenosis (AS) is currently the most common valvular heart disease in high-income countries, affecting approximately 13 million individuals worldwide^1^, and remains one of the select few cardiovascular conditions without effective medical therapy^2^. In most cases, AS is caused by calcific aortic valve disease (CAVD), a progressive fibrocalcific process that leads to aortic valve narrowing. In symptomatic end-stage disease, untreated severe AS carries a 2-year mortality rate of approximately 50%^1^. Preclinical and translational evidence support inflammation as a key mechanistic contributor to CAVD pathogenesis^3–6^, prompting consideration of anti-inflammatory treatments, such as colchicine, in preventing progression of AS^7^.

Clonal hematopoiesis of indeterminate potential (CHIP) is an age-related expansion of hematopoietic progenitor cells carrying advantageous somatic mutations in the absence of overt hematologic disease^8^. CHIP is well established as an independent risk factor for atherosclerosis, heart failure, and arrhythmias^9–11^. Mechanistically, cardiovascular risk in certain forms of CHIP is attributed to gene mutations that increase inflammasome activation, leading to increased interleukin-1 beta (IL-1β) production^12^. Accordingly, anti-inflammatory treatments such as colchicine and the IL-1β antibody canakinumab are proposed as potential targeted therapies to mitigate CHIP-attributed cardiovascular risk^13–15^.

A handful of studies have evaluated the role of somatic mosaicism in CAVD. Individuals with CHIP (particularly those with *DNMT3A* or *TET2* CHIP, the most common driver gene mutations) or with mosaic loss of the Y chromosome have increased mortality and higher rates of hospitalization after transcatheter aortic valve replacement (TAVR)^16–19^. In a meta-analysis of data from All of Us, the UK Biobank, and BioVU, Abplanalp and colleagues established an association between CHIP and incident AS, driven by the CHIP driver genes *TET2* and *ASXL1*^20^. The authors demonstrate a putative causal role for monocytes with *TET2* CHIP in the development of aortic valve calcification. The UKB results are replicated in a recent study by Wu et al, with the additional finding that *JAK2* CHIP is associated with incident AS diagnosis^21^. CHIP is also associated with progression of AS in a small, prospective study^22^. However, mechanistic insights between CHIP driver genes beyond *TET2* and risk of AS remain limited.

In the present study, we evaluate the association of CHIP driver genes with incident AS and aortic valve hemodynamics using data from the UK Biobank (UKB) and Atherosclerosis Risk in Communities (ARIC) studies to prioritize candidate CHIP driver mutations for mechanistic investigation. We then validate findings using an *in vitro* model with human primary valve interstitial cells (VICs). Lastly, we test the hypothesis that anti-inflammatory therapies modify CHIP-associated aortic valve calcification *in vitro*.

## Methods

### Study populations

The UKB is a large, population-based prospective study of more than 500,000 individuals living in the United Kingdom (UK)^23^. UKB participants aged 40-70 years were recruited between 2006-2010 at 22 assessment centers throughout the UK. The present study was conducted under UKB application number 7089. Whole-exome sequencing (WES) was performed on blood samples drawn at study enrollment. Follow-up in the cohort is conducted via linkage to UK national health records.

ARIC is a prospective cohort study of approximately 16,000 middle-aged Black and White adults recruited from four communities in the United States (Forsyth County, North Carolina; Jackson, Mississippi; suburban Minneapolis, Minnesota; Washington County, Maryland)^24^. ARIC participants were followed over seven visits occurring between 1987 and 2019. Inclusion in the present study was limited to individuals with echocardiographic data available at both visits five (V_5_: 2011-2013) and seven (V_7_: 2018-2019), totaling 1,963 individuals. WES was performed on blood samples drawn at V_5._ This study was conducted in accordance with the principles of the Declaration of Helsinki. Ethical approval was obtained from the Mass General Brigham, UKB, and ARIC. All UKB and ARIC participants provided written informed consent.

### Ascertainment of exposures, outcomes, and clinical risk factors

The occurrence of CHIP was determined using exome-sequencing data from the UKB and ARIC as previously described^10,25^ WES in the UKB was performed using the Illumina NovaSeq 6000 platform at the Regeneron Genetics Center (Tarrytown, NY)^26^. WES in ARIC was performed using the NovSeq 6000 platform (Illumina, Inc., CA). In both the UKB and ARIC, somatic variants were identified using the GATK MuTect2 tool^27^ (Broad Institute) in the Terra platform. CHIP was detected using a publicly available pipeline (https://app.terra.bio/#workspaces/terra-outreach/CHIP-Detection-Mutect2/). Mutect2 calls were filtered based on the following criteria: variants were kept if a) total depth of coverage (DP) ≥ 20, b) number of reads supporting the alternate allele (AD_Alt) ≥ 5, c) ≥1 read in both forward and reverse direction supporting the alternate allele (F1R2_Alt and F2R1_Alt), d) variant allele fraction ≥2%, and e) gnomAD allele frequency ≤0.001 (not hotspot mutations). CHIP variants that passed these criteria underwent additional curation, as described previously^28^.

The co-primary study exposures in both ARIC and UKB were the presence of any CHIP (variant allele frequency [VAF] ≥ 2%) and large CHIP (VAF ≥ 10%; CHIP clones with higher VAF are more strongly associated with cardiovascular outcomes^29^). Secondary exposures included individual CHIP driver mutations (*TET2*, *DNMT3A*, *ASXL1*, *PPM1D, TP53,* and *JAK2*) and non-*DNMT3A* CHIP, evaluated separately for VAF ≥ 2% and VAF ≥ 10%.

The primary study outcome in the UKB was incident diagnosis of AS, which was defined using a previously validated definition incorporating *International Classification of Diseases* and *Current Procedural Terminology* codes for non-rheumatic AS and aortic valve replacement^3^. Individuals with congenital heart disease, including bicuspid aortic valves, were excluded. Covariates included age (at time of sequencing), sex, body mass index (BMI), hypertension (HTN), type 2 diabetes mellitus (T2D), estimated glomerular filtration rate (eGFR), smoking status (ever versus never), and low-density lipoprotein cholesterol (LDL-C). Biometric data, including BMI, were measured by trained study staff. Smoking status was determined via touchscreen questionnaire. Clinical phenotypes (HTN, T2D, coronary artery disease [CAD]) were determined by self-report or qualifying ICD or *Office of Population Censuses and Surveys Classification of Surgical Operations and Procedures* (OPCS) codes. Relevant ICD and/or OPCS codes are presented in **Supplemental Table 1**.

The co-primary study outcomes in ARIC were V_7_ aortic valve peak velocity (V_peak_) and aortic valve mean gradient. Echocardiograms in ARIC were performed at V_5_ and V_7_ using a standardized protocol^30^. Secondary outcomes included V_peak_ and mean gradient measured at V_5_. Clinical covariates included age, sex, BMI, HTN, T2D, eGFR, smoking status (ever versus never), LDL-C, and left ventricular ejection fraction (LVEF), all of which have described in detail^31^.

### Lentiviral introduction of ASXL1 variants into human THP-1 monocytes

Human THP-1 derived macrophage like cells were cultured at 5×10^5^ cells/mL in filtered RPMI (Gibco #11875-093) media containing 0.05 µM BME (Sigma-Aldrich #63689-100ML-F), 10% FBS (Gibco #A5669501), and 1% Pen Strep (Gibco #15140-122). Human THP-1 derived macrophage like cells were spinfected (900g x 25 minutes) with 4 µg/mL polybrene (Millipore Sigma, TR-1003) and lentiviruses packed with CMV-IRES-EGFP or CMV-ASXL1_G646*-IRES-EGFP plasmids derived from the study by Balasubramani et al^32^. Transductions were verified by flow cytometry and titrated to mixtures of 10% EGFP+/90% EGFP+ cells to model clonal hematopoiesis. Following lentivirus transduction, monocytes were differentiated with 25 nM phorbol 12-myristate 13-acetate (PMA) (Thermo Scientific #356150010) for 48 hours, washed, incubated in fresh media without PMA for 24 hours, then subjected to inflammasome or media transfer experiments.

### Inflammasome activation studies

For NOD-, LRR- and pyrin domain-containing protein 3 (NLRP3) inflammasome assays, differentiated human THP-1 derived macrophage like cells were incubated with lipopolysaccharide (LPS) (1 µg/mL) (Invitrogen # tlrl-peklps) for one hour. LPS treated media was removed, then cells were incubated with 10 µM nigericin (Tocris Bioscience # 28643-80-3) for 90 minutes. Media was collected for IL-1β (R&D Systems #DY201) quantification by ELISA assay per manufacturer’s instructions. LDH activity was quantified with CyQUANT^TM^ LDH Cytotoxicity Assay (Invitrogen #CD20301). Absorbance values were measured with a Molecular Devices Spectramax 190 Microplate Reader and SoftMax Pro 7.0 software.

For AIM2 Inflammasome assays, differentiated human THP-1 derived macrophage like cells were incubated with 0.3 µg poly(dA:dT) (InvivoGen, #tlrl-patn-1) and Lipofectamine 2000 (Invitrogen, #11668019) diluted in OptiMEM (Gibco, #31985-062) for eight hours as previously described^33^. No LPS was administered. Cell media was collected for IL-1β (R&D Systems #DY201) and IL-6 (R&D Systems #DY206) with quantification by ELISA assay per manufacturer’s instructions. LDH activity was quantified with CyQUANT^TM^ LDH Cytotoxicity Assay (Invitrogen #CD20301).

Absorbance values were measured with a Molecular Devices Spectramax 190 Microplate Reader and SoftMax Pro 7.0 software.

### Media transfer experiments

Following differentiation protocol, human THP-1 derived macrophage like cells were incubated in the presence or absence of 0.3 µg poly(dA:dT) (InvivoGen, #tlrl-patn-1) and Lipofectamine 2000 (Invitrogen, #11668019) diluted in OptiMEM (Gibco, #31985-062) for eight hours (no LPS was administered). After eight hours, cells were washed with PBS, and the media was collected 24 and 48 hours later. Supernatants were pooled, cell debris was removed by centrifugation, and media were frozen for calcification studies.

### Human valvular interstitial cell isolation

Human aortic valve (AV) leaflets were collected from valve replacement surgeries for severe AS at Brigham and Women’s Hospital. This study was conducted in accordance with the principles of the Declaration of Helsinki. Ethical approval was obtained from the Brigham and Women’s Hospital (BWH IRB protocol number: 2011P001703) and all participants provided written, informed consent. The AV samples were kept on ice in DMEM culture media (Corning, 10-013-CV) and then washed in PBS three times. Human primary VICs were isolated from the AV leaflets using collagenase digestion. After cutting into 1-2 mm pieces, sections were digested using 1 mg/mL collagenase (Sigma, C5894) in DMEM at 37 °C for 10 min with gentle mixing.

Valvular endothelial cells were removed by washing with DMEM and subsequently discarded. After washing with PBS, AV pieces were further digested using 2 mg/mL collagenase type 2 (Worthington, LS004174) for 20 hours with gentle mixing. The cell suspension was filtered using 40 µm cell strainer, and isolated VICs were collected by centrifugation (1,000 rpm, 5 minutes) and plated in 75 cm^2^ culture flasks. Isolated VICs were cultured in growth media (GM) containing DMEM supplemented with 10% FBS, 1% penicillin/streptomycin (PS) (Lonza, 17-602E) in a CO_2_ incubator (37°C, 5% CO_2_) until the cells were >90% confluent. Cells were then detached using 0.05% trypsin/EDTA (Thermo Fisher Scientific, 25200056) and plated for subculture. VIC passage 4-7 were used for all experiments.

### Calcification assay of VICs

VICs were plated in 48-well plates at a density of 1 × 10^5^ cells/mL using GM. After 24 hours, VICs were cultured with a mixture of GM and THP-1 macrophage conditioned media at a ratio of 1:1. For inhibition experiments, IL-1 receptor antagonist (Anakinra, MedChemExpress, HY-108841: final concentration 1,000 µg/mL), anti-IL-6 receptor antibody (Tocilizumab, MedChemExpress, HY-P9917; final concentration 10 µg/mL), or human IgG isotype control (Invitrogen, MA5-55090; final concentration 10 µg/mL) was added to culture media. The cell culture media was changed every 3-4 days. Calcium deposition was detected using 2% Alizarin Red S staining solution (Lifeline Cell Technology, CM-0058). VICs were fixed with 10% formalin for 15 minutes and washed with distilled water. After adding Alizarin red S staining solution, cells were incubated for 30 minutes at room temperature. Excess stain was removed by washing 3 times with distilled water. Alizarin Red S staining was extracted using 5% formic acid and calcium content was quantified by absorbance at 450 nm.

### Mass spectrometry-based proteomics

Conditioned media from human THP-1 derived macrophage like cells were subjected to mass spectrometry analysis. Samples were processed in triplicate using the iST-BCT kit (PreOmics, Germany) according to the manufacturer’s instructions with the following adjustments.

Samples were digested for 1 hour at 37 °C , eluted peptides were evaporated at 45 °C until dry, and resuspended in 25 µL LC-load prior to peptide quantification. Tryptic peptides were analyzed using the quadrupole Orbitrap Exploris 480™ coupled to a Vanquish™ Neo UHPLC system (Thermo Fisher Scientific). The resulting peptides (800 ng on column) were fractionated using a dual column set-up: a PepMap™ Neo C18 Trap Cartridge, 5 μm, 300 μm X 5 mm (Thermo Fisher Scientific, Cat# 174500); and an Easy-Spray™ PepMap™ Neo C18 Column, 2 μm, 75 μm X 150 mm (Thermo Fisher Scientific, Cat# ES75150PN). The column was heated at a constant temperature of 45 °C. The gradient flow rate was 300 nL/min from 5 to 21% solvent B (95% acetonitrile /0.1% formic acid in mass spectrometry-grade water) for 50 min, 21 to 30% solvent B for 10 min, and another 15 min of a 95%-5% solvent B sawtooth wash. Solvent A was 0.1% formic acid in the water. The mass spectrometry analysis was performed using data-independent acquisition (DIA). MS1 scans were acquired in profile mode and MS2 scans in centroid mode. Spray voltage was set to 2,000 V, funnel RF level at 50, and heated capillary temperature at 275 °C. The MS1 scan range of m/z400–900 was set to 120,000 resolution. The automatic gain control (AGC) target was set to standard with a maximum injection time of 45 ms. MS2 spectra were acquired with a precursor isolation range of m/z 400−900 divided into 8 Th windows with an overlap of 1 Da. The MS2 resolution was set to 30,000 and the AGC target was set at 1000% with a maximum injection time of auto. HCD collision energy was set to 26%.

### DIA data processing

The acquired peptide spectra were analyzed using DIA-NN version 2.0 Academia. The timsTOF HT spectra were analyzed using DIA-NN 2.0 Enterprise. The spectra were queried against the same Uniprot human database (downloaded May 2025; 42,421 entries) in library-free mode. Trypsin/P was set as the digestion enzyme, allowing up to 1 missed cleavage and a minimum peptide length of 7-30 amino acids (1-4 peptide charge). N-terminal methionine excision, carbamidomethylation (+57.021 Da) of cysteine and oxidation (+15.995 Da) of methionine were considered. The precursor FDR was 1.0%. For the algorithm settings, match between runs and protein inference were selected. The quantification strategy was set to Quant UMS (high precision), and the cross-run normalization was retention time-dependent.

### Statistical analysis

All continuous variables in the UKB and ARIC were evaluated for normality using the Shapiro-Wilk test on a random subset of 5,000 individuals. Normally distributed continuous features were compared between individuals with and without CHIP using Student’s t*-*test; skewed variables were compared using the Wilcoxon rank-sum test. Categorical variables were compared between groups using chi-squared tests.

Covariates in the UKB with missingness > 2% were imputed using multiple imputation by chained equations implemented by the mice package (v3.17.0) in R. We generated five imputed datasets specific to variable type: predictive mean matching for continuous variables, logistic regression for binary variables, polytomous for unordered categorical variables, and proportional odds logistic regression for ordered variables. For each dataset, outcome and exposure variables were excluded; all other covariates (e.g., age, sex, genetic ancestry, BMI, HTN, T2D, smoking, eGFR, and CAD) were used as predictors. A single, complete dataset was then derived from multiply imputed data for downstream analysis. Hazard ratios for incident AS in UKB were calculated using Cox proportional hazards models. Proportional hazards assumptions were tested by calculating Schoenfeld residuals. Baseline adjustment for all models included age, sex, and race. Fully adjusted models additionally included BMI, HTN, T2D, eGFR, smoking status (ever versus never), and LDL-C. Prevalent conditions were assigned at UKB enrollment. Subjects not diagnosed with AS were censored at the end of follow-up or death. Statistical significance was considered as a two-sided P-value < 0.05 (for the co-primary exposures of any CHIP and large CHIP, which are highly correlated). Subgroup analyses were performed for significant associations in fully adjusted models comparing effect estimates and calculating a P-value for interaction between subgroups by CAD, HTN, sex, smoking status (ever-vs. never-smoker), and T2D.

Covariates in ARIC with missingness > 2% were imputed using the mice package (v3.17.0) in R using a similar schema as described in UKB. Linear regression was performed in ARIC to determine the association between CHIP exposures, V_peak,_ and mean AV gradient. We used the natural logarithm of V_peak_ and mean gradient to reduce skewness. Baseline adjustment for all models included age, sex, and race. Fully adjusted models additionally included BMI, HTN, T2D, eGFR, smoking status, and LDL-C. Statistical significance was considered as a two-sided P-value < 0.05 (for the co-primary exposures of any CHIP and large CHIP). As in UKB analyses, subgroup analyses were performed for significant associations in fully adjusted models comparing effect estimates and calculating a P-value for interaction between subgroups of individuals by CAD, HTN, sex, smoking status (ever-vs. never-smoker), and T2D. We also performed two sensitivity analyses in ARIC to assess significant findings: first, we removed individuals with clinical aortic stenosis (V_peak_ ≥ 2.5 m/s) and re-evaluated the association with aortic valve hemodynamics; and second, we adjusted models for left ventricular ejection fraction (LVEF).

Staining intensity for AV calcification by group and ELISA results for IL-1β and IL-6 in media of different conditions were compared using Student’s t-tests (2-tailed; paired) utilizing Prism 10 (GraphPad version 10.6.1).

Proteomics abundances from mass spectrometry were compared using empirical Bayes-moderated linear models implemented in the *limma* packag^34^. Proteins were retained for analysis if present in at least two of three technical replicates. All protein abundances were log_2_-transformed prior to running comparisons.

## Results

### Baseline characteristics and CHIP frequency in UKB and ARIC

The final study sample in the UKB consisted of 449,109 individuals with WES and without prevalent AS or congenital heart disease (**Supplemental Figure 1**). Over a median (Q1-Q3) follow-up of 13.6 (12.8-14.3) years, 4,271 (1.0%) individuals in the UKB had an incident diagnosis of AS. Baseline characteristics between those with and without incident AS, including CHIP counts, in the UKB are displayed in **Supplemental Table 2**. There were 15,515 (3.5%) individuals with any CHIP and 10,335 (2.3%) individuals with large CHIP. The most common CHIP driver mutation was *DNMT3A* (n = 9,615 individuals, 2.1%), followed by *TET2* (n = 2,228 individuals, 0.5%), and *ASXL1* (n = 1,724 individuals, 0.4%). Baseline demographics for individuals with and without CHIP in the UKB are displayed in **Table 1**. Individuals with CHIP were older at enrollment, more commonly male, and were more frequently comorbid with prevalent cardiometabolic conditions, including greater frequencies of prevalent T2D, CAD, HTN, higher BMI, and lower eGFR. We also compared individuals with non-*DNMT3A* CHIP to *DNMT3A* CHIP. Compared to *DNMT3A* CHIP, individuals with non-*DNMT3A* CHIP were more frequently male and were more frequently comorbid with cardiometabolic conditions including T2D, CAD, and HTN.

**Table 1.** Baseline demographics, clinical risk factors, and CHIP status for individuals in the UK Biobank with and without CHIP.

| Feature | No CHIP<br>N = 433,594 | Any CHIP<br>N = 15,515 | P-value | *DNMT3A*<br>CHIP<br>N = 9,615 | Non-*DNMT3A*<br>CHIP<br>N = 5,900 | P-value |
| --- | --- | --- | --- | --- | --- | --- |
| Age (mean, SD), years | 56.4 (8.1) | 60.7 (6.7) | <0.001 | 61.3 (6.6) | 60.3 (6.8) | <0.001 |
| Male sex (N, %) | 197,667 (46%) | 7,274 (47%) | 0.001 | 3,985 (41%) | 3,289 (56%) | <0.001 |
| Asian (N, %) | 9,909 (2%) | 260 (2%) | <0.001 | 156 (2%) | 104 (2%) | 0.55 |
| Black (N, %) | 6,875 (2%) | 159 (1%) | <0.001 | 116 (1%) | 43 (0.7%) | 0.005 |
| White (N, %) | 408,294 (94%) | 14,845 (96%) | <0.001 | 9,177 (95%) | 5,668 (96%) | 0.04 |
| Mixed (N, %) | 2,541 (1%) | 72 (0.5%) | 0.06 | 50 (0.5%) | 22 (0.4%) | 0.24 |
| Other (N, %) | 3,933 (1%) | 108 (0.7%) | 0.007 | 74 (0.8%) | 34 (0.6%) | 0.19 |
| **Prevalent conditions** |  |  |  |  |  |  |
| Type 2 diabetes (N, %) | 10,362 (2%) | 503 (3%) | <0.001 | 265 (3%) | 238 (4%) | <0.001 |
| eGFR (mean, SD) | 94.6 (13.1) | 91.4 (13.4) | <0.001 | 91.8 (13.1) | 90.7 (13.7) | <0.001 |
| Coronary artery disease (N, %) | 17,622 (4%) | 918 (6%) | <0.001 | 465 (5%) | 453 (8%) | <0.001 |
| BMI (mean, SD), kg/m<sup>2</sup> | 27.4 (4.8) | 27.5 (4.6) | <0.001 | 27.3 (4.6) | 27.8 (4.6) | <0.001 |
| Hypertension (N, %) | 125,024 (28%) | 5,510 (36%) | <0.001 | 3,270 (34%) | 2,240 (38%) | <0.001 |
| Ever smoking (N, %) | 193,908 (45%) | 7,997 (52%) | <0.001 | 4,798 (50%) | 3,199 (54%) | <0.001 |
| LDL-C (mean, SD), mg/dL | 138 (33.6) | 136 (34.7) | <0.001 | 138 (34.7) | 134 (34.4) | <0.001 |
| **Outcome** |  |  |  |  |  |  |
| Aortic stenosis (N, %) | 4,016 (0.9%) | 255 (2%) | <0.001 | 125 (1%) | 130 (2%) | <0.001 |

**Footnote:** Baseline characteristics in the UK Biobank comparing individuals with versus without CHIP and with *DNMT3A* CHIP versus non-*DNMT3A* CHIP. Abbreviations as follows: AS (aortic stenosis); SD (standard deviation); eGFR (estimated glomerular filtration rate); BMI (body mass index); CHIP (clonal hematopoiesis of indeterminate potential); LDL-C (low density lipoprotein cholesterol). P-values were generated by Student's t-test (for continuous, normally distributed variables), Wilcoxon rank-sum test (for continuous, skewed variables), or chi-squared tests (for categorical variables).

The ARIC analytic cohort included 1,963 individuals with genotyping and echocardiography data at both V_5_ and V_7_. Baseline demographics for ARIC individuals at both visits are displayed in **Table 2**. The majority of individuals in ARIC had subclinical CAVD with a median V_peak_ 1.3 (interquartile range 1.1-1.4) m/s at V_5_ and 1.4 (interquartile range 1.2-1.6) m/s at V_7_. There were 23 (1%) individuals at V_5_ and 81 (4%) individuals at V_7_ with at least mild AS (V_peak_ ≥ 2.5 m/s). There were 445 (23%) individuals with CHIP and 148 (8%) individuals with large CHIP in ARIC at V_5_. The most common CHIP driver mutation was *DNMT3A* (n = 234 individuals, 12%), followed by *TET2* (n = 80 individuals, 4%), and *ASXL1* (n = 25 individuals, 1%).

**Table 2.**
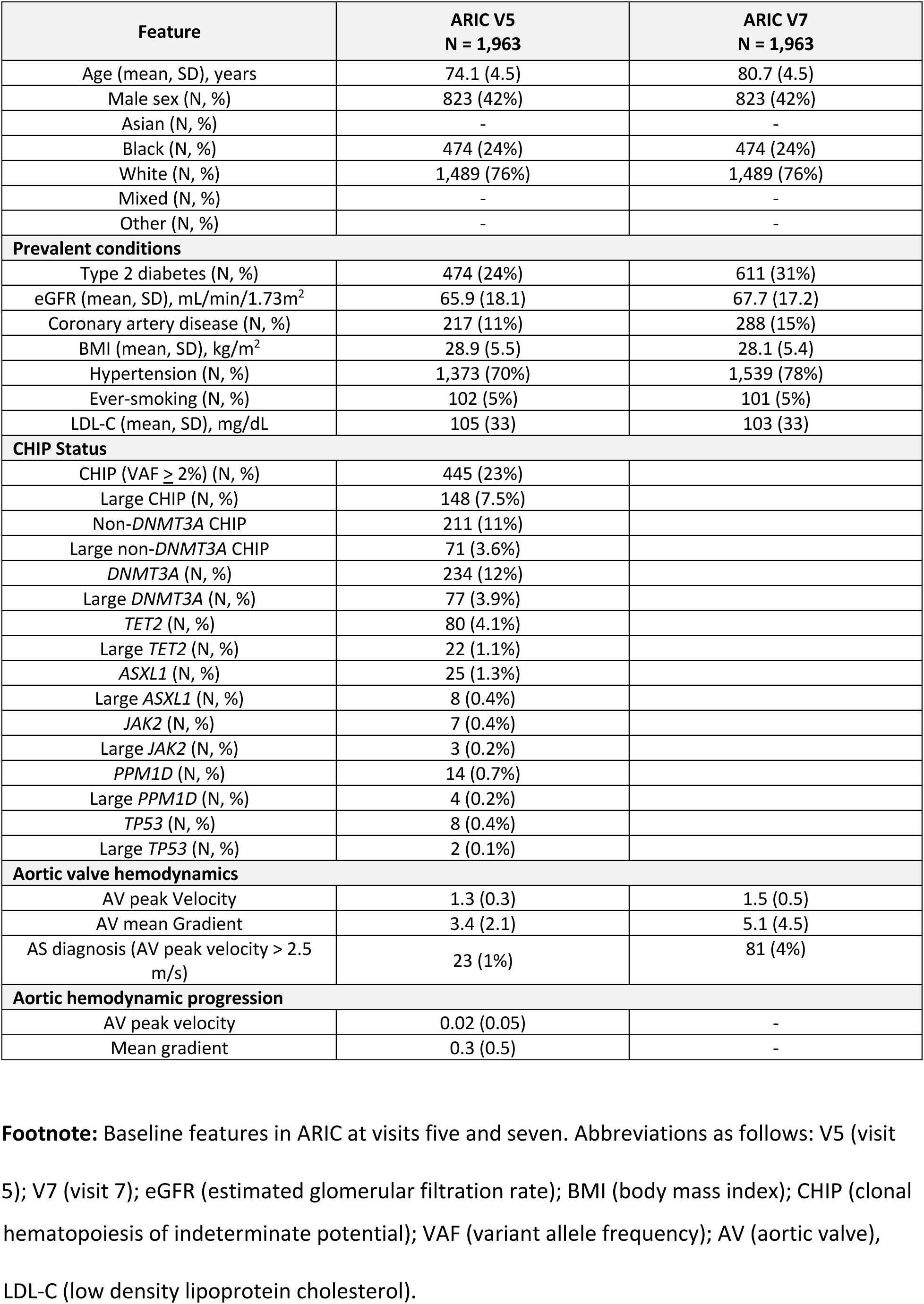
Baseline demographics, risk factors, and CHIP status for ARIC visits five and seven.

### Association of CHIP with incident AS in the UKB

In fully adjusted analysis, large CHIP versus no CHIP was significantly associated with incident diagnosis of AS in the UKB (HR 1.27, 95% confidence interval [CI] 1.09-1.47, P=0.002), whereas the association between any CHIP and AS was not significant (HR 1.12 [95% CI 0.98-1.27], P=0.09) (**Figure 1, Supplemental Table 3**). Both non-*DNMT3A* CHIP and large non-*DNMT3A* CHIP were significantly associated with incident AS (non-*DNMT3A* CHIP HR: 1.29 [95% CI 1.08-1.55], P=0.005; large non-*DNMT3A* CHIP HR: 1.39 [95% CI 1.14-1.69], P=0.001). Among individual driver gene mutations, we observed associations of *TET2*, large *TET2*, *ASXL1*, large *ASXL1*, *JAK2*, large *JAK2*, large *PPM1D*, and *TP53* with incident AS, with effect estimates similar to prior reports^20,21^. The numerically largest magnitude of association was seen for *JAK2* CHIP (HR: 2.75 [95% CI 1.31-5.77]; P=0.008), followed by *TP53* CHIP (HR: 2.22 [95% CI 1.14-3.73], P=0.02). In subgroup analyses, there were no significant interactions between CHIP exposures and association with incident AS across subgroups stratified by sex, HTN, T2D, or ever-smoking status (**Supplemental Table 4**). Large *ASXL1* CHIP, which is strongly linked to history of smoking, remained significantly associated with AS among never-smokers (HR: 2.20 [95% CI 1.28-3.80]; P=0.005; **Supplemental Table 4**).

**Figure 1.**
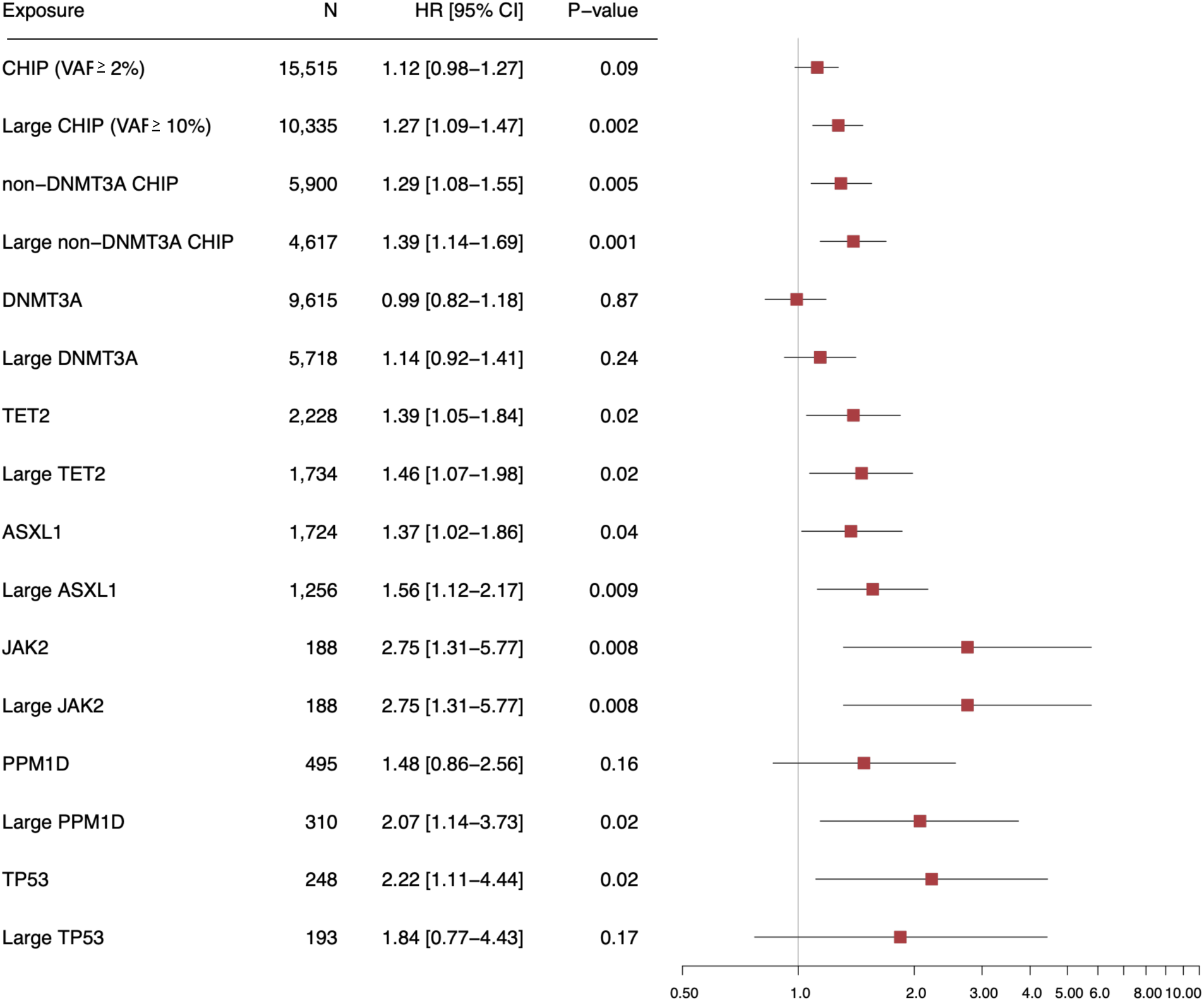
Multivariable-adjusted associations of CHIP and key CHIP subtypes with incident aortic stenosis in the UK Biobank Forest plot of hazard ratios for risk of incident aortic stenosis by CHIP status in the UK Biobank. Hazard ratios and 95% confidence intervals are shown for each CHIP exposure in a fully adjusted analysis accounting for age, sex, and prevalent hypertension, diabetes, estimated glomerular filtration rate, and smoking status. Abbreviations as follows: HR (hazard ratio); CI (confidence interval); CHIP (clonal hematopoiesis of indeterminate potential); VAF (variant allele frequency); AV (aortic valve).

### Association of CHIP with aortic valve hemodynamics and calcification in ARIC

To prioritize gene driver candidates for mechanistic study, we additionally evaluated the association between individual gene driver mutations and subsequent aortic valve hemodynamics in ARIC. In fully adjusted analyses among 1,963 ARIC study participants at V_7_, neither CHIP nor large CHIP were associated with V_peak_ or mean gradient (CHIP: AV V_peak_, beta 0.05 [95% CI −0.06-0.15] for those with versus without CHIP, outcome modeled on the natural log scale, P=0.39; AV mean gradient, beta 0.04, [95% CI −0.07-0.14], P=0.47; large CHIP: AV V_peak_, beta 0.02 [95% CI −0.15-0.19], P=0.85; AV mean gradient, beta 0.01 [95% CI −0.16-0.18], P=0.88).

In gene-specific analyses, both *ASXL1* CHIP and large *ASXL1* CHIP were nominally significantly associated with unfavorable aortic valve hemodynamic parameters at V_7_ (*ASXL1* CHIP: V_peak_, beta 0.44 [95% CI 0.05-0.84], P=0.03; AV mean gradient, beta 0.45 [95% CI 0.06-0.85], P=0.02; large *ASXL1* CHIP: V_peak_, beta 0.74 [95% CI 0.06-1.42], P= 0.03; mean gradient, beta 0.82 [95% CI 0.15-1.49], P=0.02) (**Table 3**). When we excluded individuals with mild or more severe AS from the analysis (V_peak_ ≥ 2.5 m/s, n = 81 individuals removed), large *ASXL1* CHIP remained nominally significantly associated with higher aortic valve gradients and velocities (**Supplemental Table 5**). Both *ASXL1* CHIP and large *ASXL1* CHIP maintained nominally significant associations with aortic valve hemodynamics after additional adjustment for LVEF (**Supplemental Table 6**). There were no significant interactions between CHIP exposures and association with aortic valve hemodynamics among subgroups stratified by CAD, HTN, sex, ever-smoking status, or T2D (**Supplemental Table 7**).

**Table 3.** Associations between CHIP exposures and aortic valve hemodynamics in ARIC.

| Exposure | AV peak velocity<br>log(m/s) |  | AV mean gradient<br>log(mmHg) |  |
| --- | --- | --- | --- | --- |
|  | Beta (95% CI) | P-Value | Beta (95% CI) | P-Value |
| CHIP (n=445) | 0.05 (-0.05-0.15) | 0.39 | 0.04 (-0.07-0.15) | 0.44 |
| Large CHIP (n=148) | 0.02 (-0.15-0.19) | 0.85 | 0.01 (-0.16-0.18) | 0.88 |
| non-*DNMT3A* (n=211) | 0.12 (-0.02-0.27) | 0.10 | 0.11 (-0.04-0.25) | 0.15 |
| Large non-*DNMT3A* (n=71) | 0.01 (-0.22-0.25) | 0.92 | 0.02 (-0.21-0.26) | 0.85 |
| *DNMT3A* (n=234) | -0.03 (-0.17-0.12) | 0.73 | -0.02 (-0.16-0.12) | 0.77 |
| Large *DNMT3A* (n=77) | 0.02 (-0.22-0.25) | 0.89 | 0.00 (-0.23-0.23) | 0.99 |
| *TET2* (n=80) | 0.06 (-0.16-0.29) | 0.58 | 0.07 (-0.16-0.29) | 0.56 |
| Large *TET2* (n=22) | -0.20 (-0.61-0.22) | 0.36 | -0.16 (-0.58-0.25) | 0.45 |
| *ASXL1* (n=25) | 0.44 (0.05-0.84) | **0.03** | 0.45 (0.06-0.85) | **0.02** |
| Large *ASXL1* (n=8) | 0.74 (0.06-1.42) | **0.03** | 0.82 (0.15-1.49) | **0.02** |
| *JAK2* (n=7) | 0.04 (-0.75-0.82) | 0.93 | 0.07 (-0.71-0.84) | 0.87 |
| Large *JAK2* (n=3) | -0.13 (-1.30-0.61) | 0.82 | -0.16 (-1.26-0.93) | 0.77 |
| *PPM1D* (n=14) | 0.14 (-0.39-0.67) | 0.60 | 0.11 (-0.42-0.64) | 0.68 |
| Large *PPM1D* (n=4) | -0.34 (-1.30-0.61) | 0.48 | -0.40 (-1.35-0.55) | 0.41 |
| *TP53* (n=8) | 0.32 (-0.40-1.05) | 0.39 | 0.36 (-0.36-1.08) | 0.32 |
| Large *TP53* (n=2) | -0.21 (-1.56-1.14) | 0.76 | -0.26 (-1.60-1.08) | 0.71 |

**Footnote:** Associations between CHIP exposures and aortic valve hemodynamics. Betas and 95% confidence intervals are shown for each CHIP exposure in fully adjusted analyses accounting for age, sex, prevalent hypertension, diabetes, estimated glomerular filtration rate, and smoking status. Abbreviations as follows: CI (confidence interval); SE (standard error); AV (aortic valve); CHIP (clonal hematopoiesis of indeterminate potential); VAF (variant allele frequency); AV (aortic valve). Missing aortic valve Agatston score values are due to inadequate numbers of driver mutations (e.g., 0) in either the case or control condition. P-values were generated by Student's t-test (for continuous, normally distributed variables), Wilcoxon rank-sum test (for continuous, skewed variables), or chi-squared tests (for categorical variables).

*Human ASXL1-mutant THP-1 derived macrophage like cells increase calcification of VICs* Because *ASXL1* was the only CHIP driver mutation associated with both incident AS in UKB and adverse aortic valve hemodynamics in ARIC, we prioritized *ASXL1* for mechanistic investigation. We first generated the *ASXL1* mutant in the commonly used human THP-1 derived macrophage like cell line via lentiviral transduction. Mechanistic studies focused on macrophages because CAVD is characterized by macrophage infiltration and inflammatory signaling^35,36^ and prior studies implicate macrophages as effector cells by which CHIP promotes cardiovascular disease^11^. *ASXL1* CHIP is hypothesized to increase cardiovascular risk by heightening the inflammatory response to AIM2 agonism^37^. Therefore, we examined inflammasome activation in human THP-1 derived macrophage like cells harboring an *ASXL1* mutation. We evaluated IL-1β secretion and lactate dehydrogenase (LDH) activity in media from both *ASXL1*-mutant and *ASXL1-*normal cells in conditions of no inflammasome activation (baseline), AIM2 inflammasome activation with poly(dA:dT), and NLRP3 inflammasome activation with lipopolysaccharide (LPS) and nigicerin. At baseline IL-1β was not detected in the media, however following AIM2 inflammasome stimulation by poly(dA:dT) treatment both IL-1β and LDH activity increased significantly in media from *ASXL1*-mutant THP-1 derived macrophage like cells, compared with media from *ASXL1*-normal, poly(dA:dT) treated cells (P-value IL-1β P<0.001, LDH activity P<0.001) (**Figure 2A**, **Supplemental Figure 2**). In contrast, there was no significant difference across genotypes for the baseline or NLRP3-activated conditions (**Figure 2A**, **Supplemental Figure 2**).

**Figure 2.**
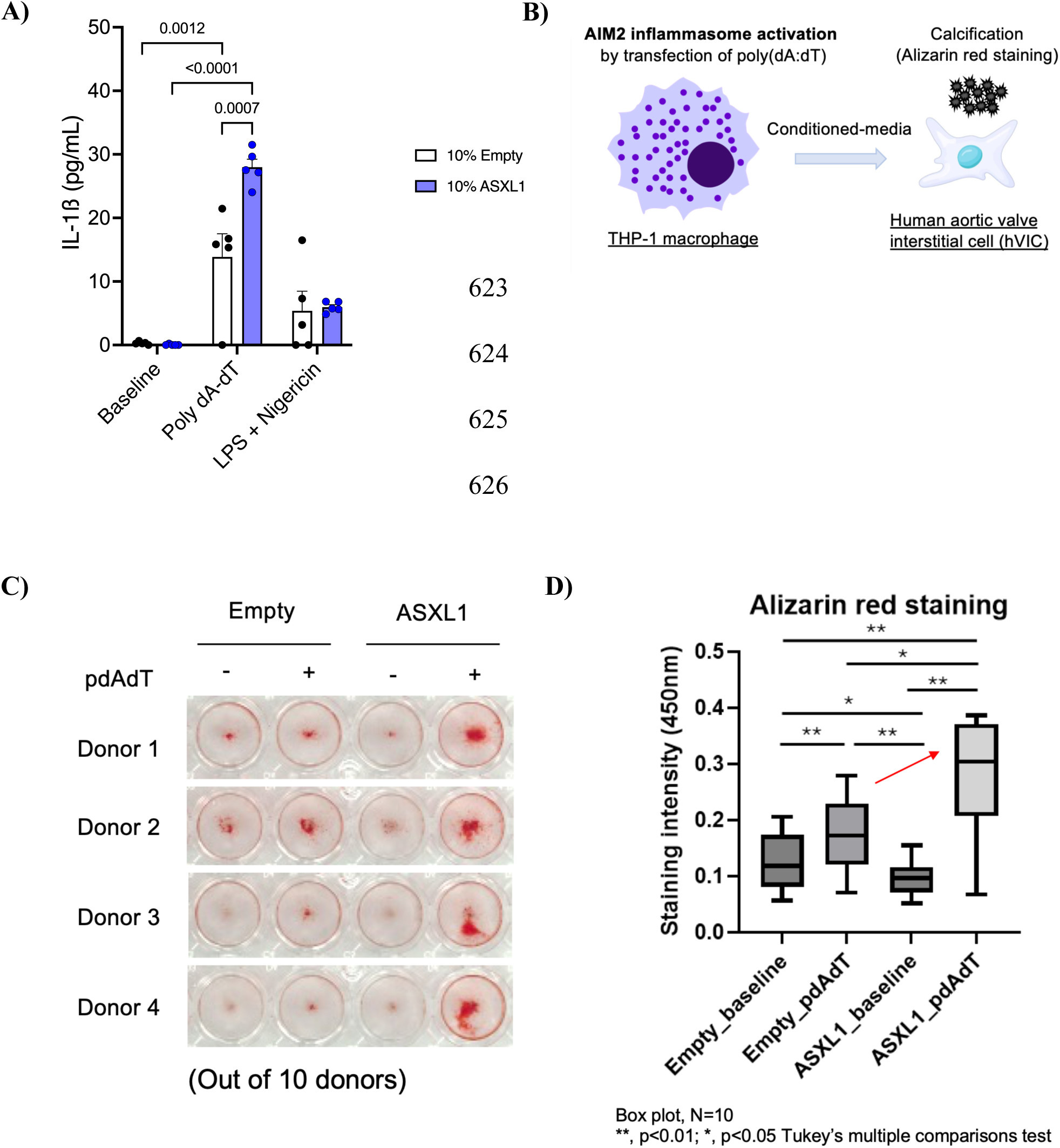
Conditioned-media of *ASXL1* mutant-overexpressed THP-1 like cells activated by poly(dA:dT) increases human valve interstitial cell calcification A: IL-1β concentrations in media from *ASXL1-*normal THP-1-derived 631 macrophage like cells (white) and *ASXL1-*mutant THP-1 macrophages (blue) across baseline (control), poly(dA:dT) treatment to activate the AIM2 inflammasome, and lipopolysaccharide (LPS) + nigicerin to activate the NOD-, LRR- and pyrin domain-containing protein 3 inflammasome. B: Schematic of experimental design. Media from THP-1 macrophages in four experimental conditions (+/- lentiviral *ASXL1* mutation, +/- AIM2 inflammasome activation by poly(dA:dT)) was added to hVICs culture for 13-17 days. Cells were then stained with Alizarin Red S to quantify calcification. C: Representative images of hVICs stained with Alizarin Red S under all four experimental conditions. Darker red staining implicates greater calcification. D: Box plot comparison of Alizarin Red S staining intensities across all four experimental conditions. Box plots show the distribution from minimum to maximum values. The box represents quartile 1 and quartile 3, with the line inside the box indicating the median. Whiskers extend to the minimum and maximum data points. N=10 donors per each condition. **, p<0.01; *, p<0.05. Tukey’s multiple comparison test.

Clonal hematopoiesis mutations are confined to the hematopoietic compartment, therefore the role of *ASXL1* mutant cells on aortic valve calcification is likely due to paracrine signaling between macrophages and VICs. We cultured VICs in media from control or *ASXL1* mutant THP-1 derived macrophage like cells (**Figure 2B**) treated with or without AIM2 inflammasome agonists poly(dA:dT) (10 donors per each condition). We then stained VIC cultures from each experimental condition with Alizarin Red S and quantified calcification. Among VICs treated with AIM2 inflammasome-activated, poly(dA:dT) treated THP-1 cell media, we found that hVICs cultured in media from the THP-1 *ASXL1* mutant condition had significantly higher staining intensity by Alizarin Red S (indicating greater calcification) than those grown in media from the THP-1 *ASXL1* normal condition (**Figure 2C,D**).

In conditioned media, we similarly compared IL-1β and IL-6 levels at baseline and experimental conditions. While not directly activated by the AIM2 inflammasome, IL-6 may be indirectly elevated as a downstream consequence of the inflammatory response, is a well-established risk factor for CAVD, and a previous report indicated that IL-6 is increased in serum from patients with *ASXL1* CHIP^12^. We found, similar to our previous analysis, that IL-1β was undetectable at baseline for both *ASXL1*-normal and *ASXL1*-mutant human THP-1 derived macrophage like cell conditions, but increased substantially with poly(dA:dT) treatment to 84.7 (SD 9.5) pg/mL in media from *ASXL1*-normal THP-1 derived macrophage like cells and 128.5 (SD 3.7) pg/mL in media from *ASXL1*-mutant THP-1 derived macrophage like cells (**Supplemental Figure 3A**). IL-6 concentrations were 1.13 (SD 0.20) pg/mL in media from *ASXL1-*normal THP-1 derived macrophage like cells and 1.56 (SD 0.36) pg/mL in media from *ASXL1*-mutant THP-1 derived macrophage like cells (P for difference 0.03), and increased 2.1-fold from baseline after poly(dA:dT) treatment in media from *ASXL1* normal THP-1 derived macrophage like cells and 3.1-fold in media from *ASXL1*-mutant THP-1 derived macrophage like cells (**Supplemental Figure 3B**).

### Mass spectrometry proteomics characterizes an inflammatory ASXL1-mutant THP-1 macrophage secretome

We performed mass spectrometry proteomics of media from *ASXL1*-mutant, poly(dA:dT) activated human THP-1 derived macrophage like cells and compared protein abundances with results from *ASXL1* normal, poly(dA:dT) activated THP-1 derived macrophage like cells. Mass spectrometry identified 1,108 unique proteins in the media across all conditions. In comparing protein results from the *ASXL1*-mutant, poly(dA:dT) activated condition relative to the *ASXL1* normal, poly(dA:dT) activated condition, there were 23 proteins at least nominally significantly different, of which 18 were present in greater abundance in the *ASXL1*-mutant condition, including inflammasome-related proteins like Cathepsin D and IL-1β (**Supplemental Table 8**) as well as several related chemokines such as matrix metalloproteinase 9 (MMP9) and C-X-C motif chemokine ligand 11 (CXCL11). Pathway analysis of these 18 proteins clarified an enrichment in pro-inflammatory pathways including cytokine-cytokine receptor interactions (KEGG), neutrophil degranulation, and the innate immune system (Reactome) (**Supplemental Table 9**).

### Selective interleukin receptor blockade reduces calcification of hVICs in vitro

To test the hypothesis that IL-1β and/or IL-6 mediates the observed increase in hVIC calcification, we treated media from *ASXL1*-mutant THP-1 derived macrophage like cells stimulated with poly(dA:dT) with anakinra, an IL-1 receptor antagonist, IgG, or tocilizumab, a monoclonal antibody targeting the IL-6 receptor. As previously, we found a significantly higher staining intensity by Alizarin Red S (indicating greater calcification) when comparing VICs treated with poly(dA:dT) alone versus no poly(dA:dT). VICs treated with poly(dA:dT) and either anakinra or tocilizumab exhibited significantly decreased calcification compared to poly(dA:dT) alone or IgG control (**Figure 3A,B**).

**Figure 3.**
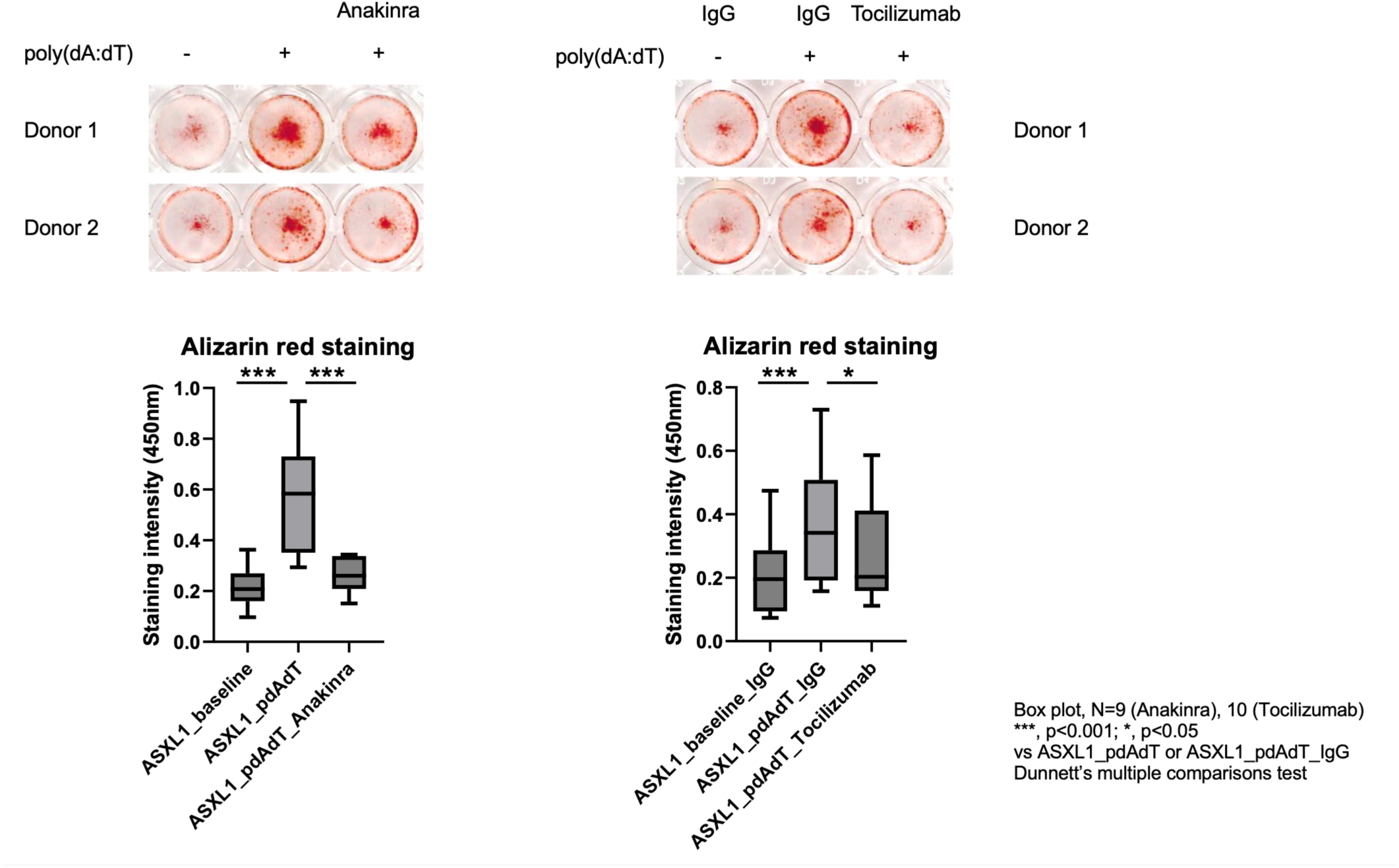
Treatment of hVICs with anakinra or tocilizumab reduces aortic valve calcification *in vitro*. Top: Representative images of VICs stained with Alizarin Red S under all experimental conditions. Darker red staining implicates greater calcification. Bottom: Box plot comparison of Alizarin Red S staining intensities across all experimental conditions. Box plots show the distribution from minimum to maximum values. The box represents the interquartile range (25th to 75th percentiles), with the line inside the box indicating the median. Whiskers extend to the minimum and maximum data points. N=9 donors (anakinra) and N=10 donors (tocilizumab). **, p<0.01; *, p<0.05. Tukey’s multiple comparison test.

## Discussion

Here, we present evidence for an association between *ASXL1* CHIP and CAVD in two large cohorts and validate this finding experimentally using human aortic valvular interstitial cells. In the UKB, gene-specific analyses indicated that the association of CHIP with AS was primarily driven by non-*DNMT3A* CHIP driver genes, including *TET2, ASXL1*, and *JAK2,* as observed in prior studies^20,21^. In ARIC, only *ASXL1* CHIP was associated with altered aortic valve hemodynamics.

We therefore focused mechanistic work on *ASXL1* CHIP. In an *in vitro* model, media from activated *ASXL1*-mutant human macrophages had elevated concentrations of the inflammatory cytokines IL-1β and IL-6 and promoted accelerated calcification of hVICs. Treatment of this media with either anakinra or tocilizumab mitigated calcification of VICs. Taken together, these data identify *ASXL1* as a CHIP driver mutation consistently associated with CAVD across complementary human cohorts and identify a plausible inflammatory pathway through which *ASXL1*-mutant myeloid cells may promote valvular calcification. These data also represent the first evidence demonstrating that human macrophage-like cells carrying *ASXL1* mutations exhibit increased inflammasome activation.

In ARIC, *ASXL1* and large *ASXL1* CHIP were independently associated with unfavorably altered aortic valve hemodynamic parameters, providing further support for the association of *ASXL1* CHIP with clinical AS diagnoses observed in the UKB in both our study and by Abplanalp et al.^20^ *ASXL1* encodes a scaffolding protein that acts as a transcriptional regulator^38^. *ASXL1*-mutant murine macrophages are observed to manifest a strong inflammatory response to AIM2 agonism^37^, implicating the AIM2 inflammasome as a primary mediator of inflammation and thereby disease risk. Accordingly, we found that media from *ASXL1*-mutant macrophages treated with poly(dA:dT), a synthetic double-stranded DNA molecule that can activate the AIM2 inflammasome, accelerated hVIC calcification *in vitro* when compared with AIM2-activated,

*ASXL1*-normal cell media. We observed a greater proportional change in IL-1β than in IL-6 following AIM2 activation, as expected given that the AIM2 inflammasome directly activates IL-1β. However, IL-6, which was also elevated, may be a relevant effector, as it is well recognized to promote VIC mineralization^39^, and *IL6* knockdown can reduce mineralization of VICs *in vitro*^40^. Accordingly, we observed that blockade of either IL-1β or IL-6 significantly mitigated VIC calcification, suggesting a potential role for both cytokines in mediating the association between *ASXL1* CHIP and CAVD.

Consistent with recent findings^20^, we observed that non-*DNMT3A* CHIP driver genes drove the association between CHIP and incident AS. *DNMT3A* CHIP has historically weaker associations with atherosclerosis than other CHIP driver genes in human populations^41^. While *in vitro* experimental models suggest *DNMT3A*, *TET2*, and *ASXL1* CHIP share some convergent pro-inflammatory effects in mice^42,43^, studies in human populations suggest that blockade of inflammatory pathways differentially impacts cardiovascular risk in individuals with *DNMT3A* versus non*-DNMT3A* CHIP. For example, a retrospective evaluation of the Canakinumab Anti-inflammatory Thrombosis Outcome Study (CANTOS) trial found that treatment with canakinumab (antibody targeting IL-1β) lowered risk of secondary cardiovascular events among individuals with *TET2* but not *DNMT3A* CHIP^13^. One potential explanation for this difference is the observation that myeloid cells with *TET2* but not *DNMT3A* variants exhibited elevated IL-1β levels after LPS treatment^44^. Moreover, murine experiments and analyses of the Low-Dose Colchicine 2 trial (LoDoCo2) suggest that low-dose colchicine may attenuate the risk associated with *TET2*, but not *DNMT3A*, CHIP^15,45^. Our data further support the use of either IL-1 or IL-6 blockade to reduce *ASXL1*-mediated aortic valve calcification.

Mass spectrometry proteomics clarified that the secretome from *ASXL1*-mutant, poly(dA:dT) activated macrophages is enriched in inflammatory proteins. In addition to observing relative enrichment in IL-1β, we found increased abundances of MMP9, CXCL11, and pro-platelet basic protein (PPBP), all of which have recognized, pro-inflammatory biological roles. MMP9, for example, may act upstream of IL-1β by processing the precursor IL-1β protein into a biologically active form^46^. MMP9 is more abundant in human valves from individuals with CAVD than in non-calcified valves^47^. CXCL11 is a potent chemotactic cytokine that functions to facilitate leukocyte migration^48^. PPBP is a related protein in the CXC family that, like CXC chemokines, facilitates leukocyte migration and has been associated with an increased risk of atherosclerotic cardiovascular disease^49,50^. Together, these proteins may represent additional potential targets beyond IL-1β to mitigate *ASXL1*-mediated aortic valve calcification.

The work has certain limitations. AS diagnoses in the UKB were ascertained using a claims-based phenotype. Importantly, our results in the UKB showed slightly different effect estimates than those in Abplanalp and colleagues’ or Wu and colleagues’ analyses. We used a previously validated claims-based definition for AS, with a sensitivity of 0.99 and specificity of 0.81 after manual chart review in the VA health system^3^, and, unlike these recent studies, excluded individuals with congenital heart disease. The majority of individuals in both UKB and ARIC were most similar to a European genetic reference panel, which may limit generalizability of findings to other populations. Finally, AS events are somewhat uncommon and occur later in life relative to other cardiovascular conditions like CAD or heart failure, which may limit power to detect associations in less common driver genes.

In summary, integrating complementary human cohort analyses with mechanistic studies, we identify *ASXL1* as a CHIP driver mutation consistently associated with CAVD. The convergence of findings from the UKB and ARIC prioritized *ASXL1* for mechanistic investigation, which implicated *AIM2*-mediated inflammatory signaling in human *ASXL1*-mutant macrophages and demonstrated that downstream IL-1 and IL-6 receptor blockade attenuated macrophage-mediated valvular interstitial cell calcification *in vitro*. Together, these findings nominate *ASXL1*-mutant CHIP as an inflammatory driver of valvular calcification and identify IL-1 and IL-6 signaling as candidate therapeutic pathways for future investigation in CAVD.

## Data Availability

All data produced in the present study are available upon reasonable request to the authors

