## Supplemental Figures for "*ASXL1-*Mutant Clonal Hematopoiesis is Associated with Calcific Aortic Valve Disease and Promotes Valvular Calcification *In Vitro*"

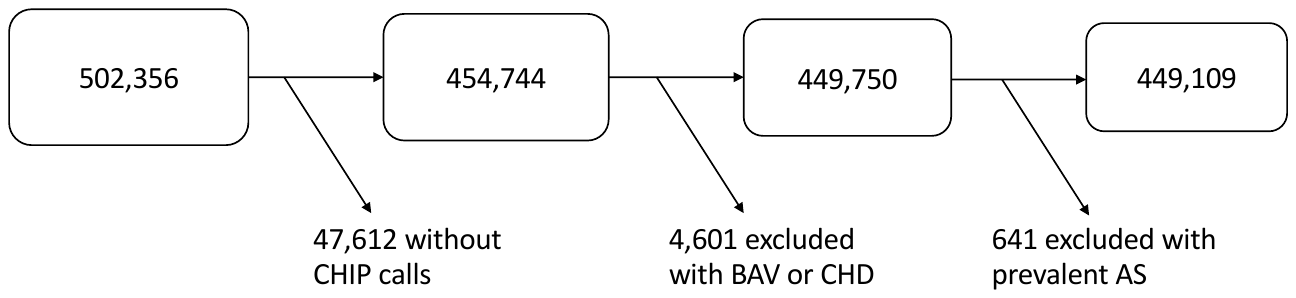
**Supplemental Figure 1:** Flow chart of individuals included in UK Biobank analysis of CHIP and incident aortic stenosis.

**Legend:** Flow chart of individuals meeting eligibility criteria for inclusion in the UK Biobank cohort of CHIP exposures and incident aortic stenosis. Baseline UK biobank population (left) included 502,356. CHIP = clonal hematopoiesis of indeterminate potential; BAV = bicuspid aortic valve; CHD = congenital heart disease; AS = aortic stenosis.

**Supplemental Figure 2:** LDH activity of media from THP-1 macrophages across experimental conditions.

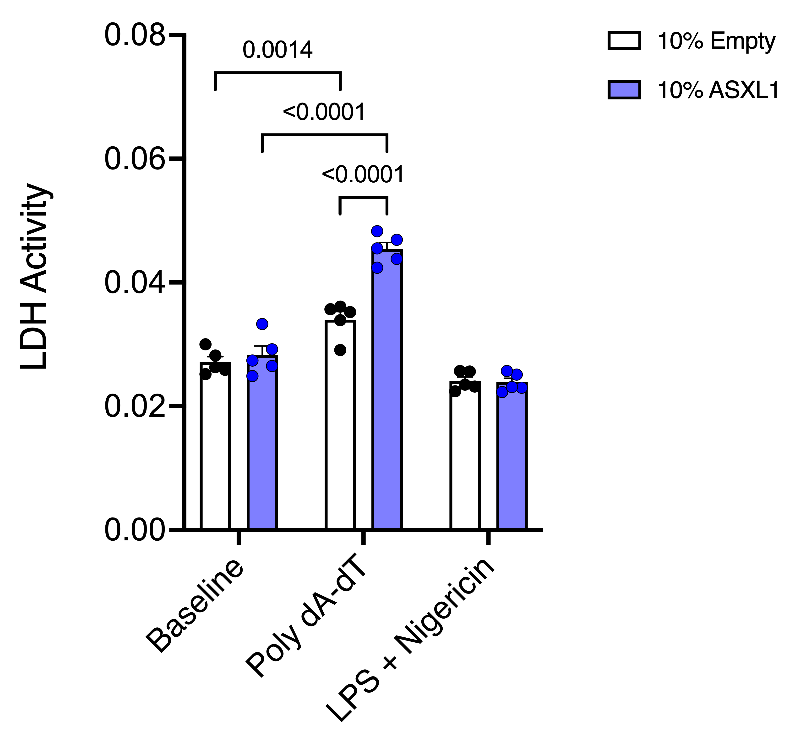

**Legend:** LDH activity in media from *ASXL1-*normal macrophages (white) and *ASXL1-*mutant THP-1 macrophages (blue) and across baseline (control), poly(dA:dT) treatment to activate the AIM2 inflammasome, and lipopolysaccharide (LPS) + nigicerin to activate the NOD-, LRR- and pyrin domain-containing protein 3 inflammasome. LDH = lactate dehydrogenase. N=5 biological replicates per condition. P-values were derived from Student’s t-tests (2-tailed; paired).

**Supplemental Figure 3:** ELISA quantification of (A) interleukin-1β and (B) interleukin-6 concentrations from media of THP-1 derived macrophage like cells across experimental conditions.

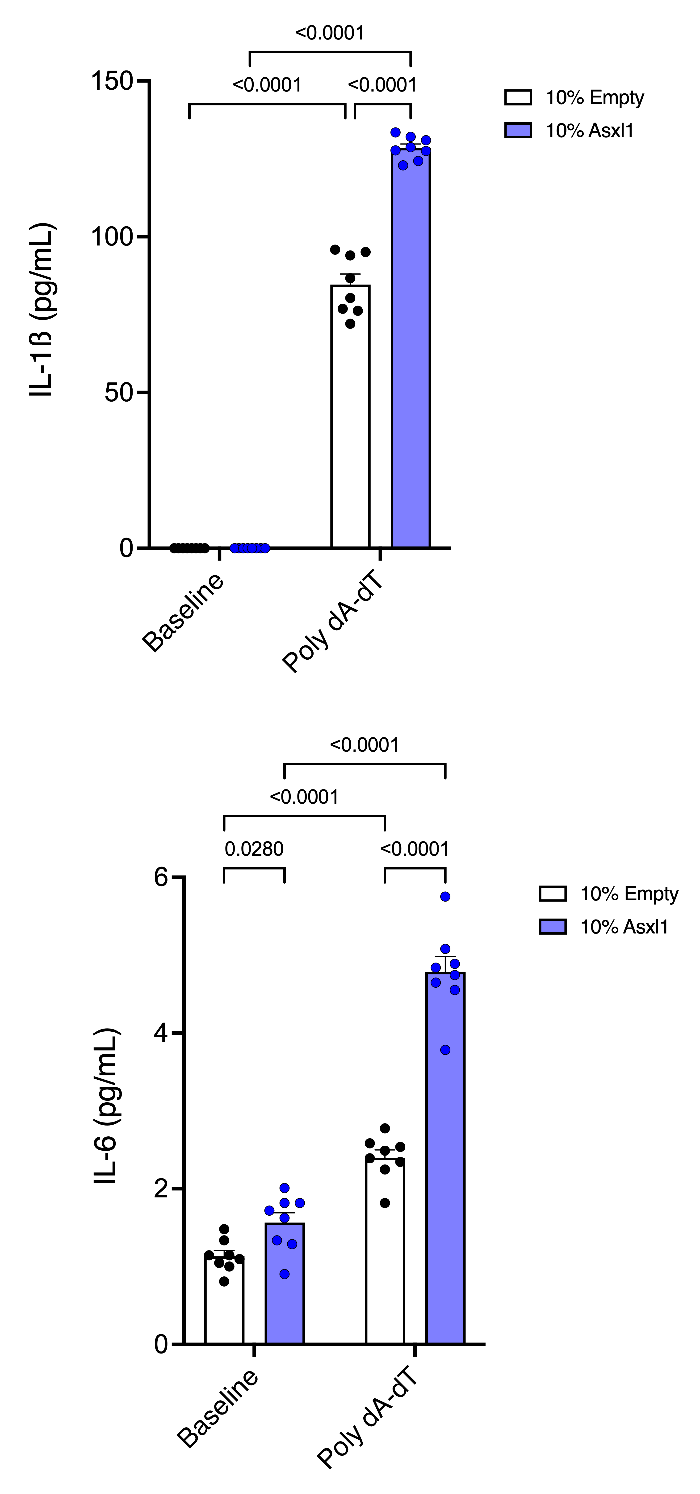

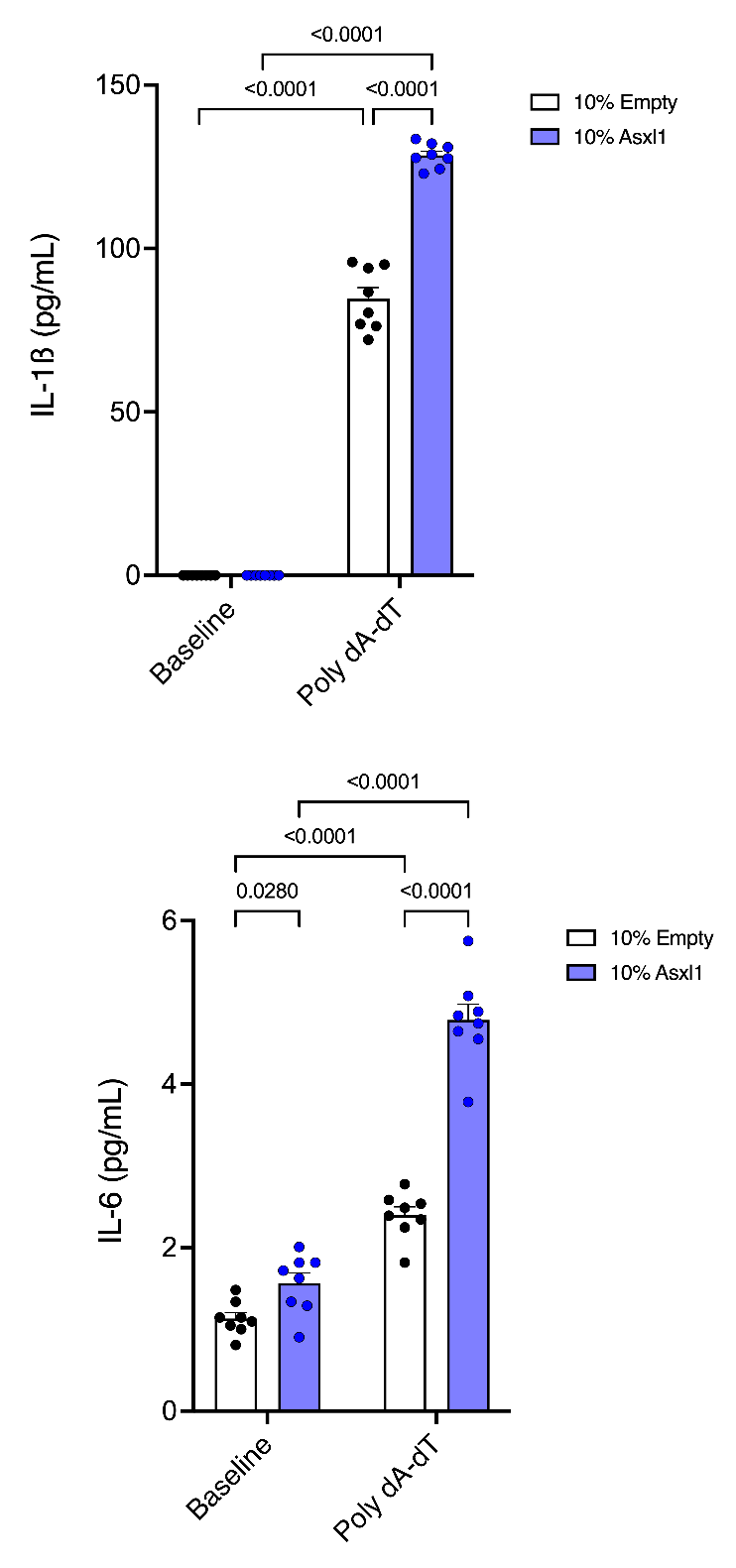

**A)**

**B)**

**Legend:** ELISA results for quantification of inflammatory cytokines comparing cells treated by an empty vector (white) and with *ASXL1* mutation (blue). A: Interleukin-1β media concentrations comparing THP-1 derived macrophage like cells with and without poly(dA:dT). B: Interleukin-6 media concentrations comparing THP-1 derived macrophage like cells with and without poly(dA:dT) treatment. IL-1β = interleukin-1 beta; IL-6 = interleukin-6. N=8 biological replicates per condition. P-values were derived from Student’s t-tests (2-tailed; paired).
